# Intervention strategies to improve contraceptive access for uninsured and underinsured women in the United States: A scoping review protocol

**DOI:** 10.1101/2025.07.03.25330797

**Authors:** Bless-me Ajani, Esther Osime, Mustapha Aliyu Muhammad, Ifedolapo Aderibigbe, Kate Beatty

## Abstract

Access to contraception is an important public health issue in the United States (U.S.), as it plays a fundamental role in preventing unintended pregnancies and improving maternal and child health outcomes. However, significant disparities in access persist in the United States, particularly among uninsured and underinsured women. These women with no or limited health insurance often experience multiple barriers, including cost, limited service availability, inadequate information, lack of access to the full range of contraceptive methods and, leading to unmet needs for contraception. The objective of this scoping review is to map evidence of intervention strategies aimed at improving access to contraceptive services for uninsured and underinsured women in the United States. The review will follow Arksey and O’Malley’s 2005 five-stage scoping review framework. We will conduct a comprehensive literature search of publications from January 2005 to June 2025 on electronic databases such as PubMed, Scopus, CINAHL, Embase, and Web of Science, as well as ScienceDirect, EMBASE, CINAHL, and PsycINFO. The PCC (Population, Concept, Context) criteria will guide study selection, focusing on interventions targeting uninsured and underinsured women in the U.S. that aim to improve contraceptive access. Two reviewers will independently screen the title, abstract and full text. We will use the TIDieR (Template for Intervention Description and Replication) checklist for data extraction to ensure comprehensive reporting of interventions. We will use narrative synthesis to analyze and summarize findings numerically and thematically, guided by Andersen’s Behavioral Model of Health Services Use framework. This scoping review will provide a comprehensive overview of intervention efforts aimed at improving access to contraception among uninsured and underinsured women in the U.S. We expect to identify strategies, programs, and policy-related interventions among this population. Based on the findings from this review, recommendations will be made for future research, program development, and policy.

**Protocol Registration:** Review registration Opens Science Framework: https://doi.org/10.17605/OSF.IO/9G6DN

## Introduction

Access to contraception is an important public health issue in the United States (U.S.), as it plays a fundamental role in preventing unintended pregnancies and improving maternal and child health outcomes [1]. Rigorous evidence synthesis is, therefore, essential to guide policy and program design. Nationally, nearly 45% of all pregnancies are unintended, with the highest rates among low-income and uninsured women [2]. Approximately 21% of non-pregnant women in the U.S. report unmet contraceptive needs [3].

Currently, about 9.3 million women of reproductive age lack health insurance, with Black, Native American, and Hispanic populations bearing the highest burden [4,5]. Women without insurance face multiple barriers, including inadequate information, limited service availability, and financial constraints, increasing their risks for unintended pregnancies and maternal morbidity or mortality [6–8]. Additionally, over 19 million women reside in contraceptive deserts, where access to family planning providers, health centers, and the full range of birth control methods are severely limited [9,10].

While 65% of reproductive-age women used contraception in 2019, many still face affordability challenges, particularly for long-acting reversible contraceptives (LARCs), which remain cost-prohibitive without insurance [11,12]. Evidence from a study on unfulfilled contraceptive preferences due to cost using the 2015-2019 National Surveys of Family Growth data revealed that 39% of low-income non-users reported they would use contraception if it were affordable [13].

In addition, evidence from the Guttmacher Institute revealed that publicly funded family planning services met only 45% of contraceptive needs in 2016.14 In the U.S., health insurance coverage is key in addressing these needs, as women with private or public insurance reported significantly lower levels of unmet contraceptive needs [13]. Despite the direct link between insurance coverage and improved access to contraceptive services, publicly funded family planning programs have only partly addressed this gap. This has led to disparities in contraceptive access, especially for low-income women with no or inadequate health insurance coverage.

### Rationale for a scoping review

Unintended pregnancy rates remain high in the U.S.; as of 2019, the rate stood at 35.7 per 1,000 women, resulting in about 886,000 abortions [14,15], reinforcing the need to identify effective, scalable interventions that address these barriers. To our knowledge, there is no synthesis of the literature or comprehensive review of evidence on interventions aimed explicitly at reaching low-income, uninsured and underinsured women in the U.S., leaving a gap in understanding how best to address the challenges faced by these vulnerable groups in accessing contraceptive services. Therefore, this scoping review seeks to map evidence of intervention strategies aimed at improving access to contraceptive services for uninsured and underinsured women in the United States. A scoping review is a suitable method for this topic because the literature spans diverse study designs, settings, and outcomes; an inclusive mapping exercise is warranted. Scoping methodology facilitates the identification of evidence gaps and emerging intervention types without restricting to effectiveness trials. Findings will provide an evidence-informed foundation to inform future research, systematic reviews, program evaluations, and policy development.

## Methods and Analysis

This scoping review will be conducted following the methodological framework that was proposed by Arksey and O’Malley [16]. The framework includes five step-by-step stages: i) Identifying the research question, ii) Identifying relevant studies, iii) Study selection, iv) Charting the data, and v) Collating, summarizing, and reporting the results.

### Theoretical/Conceptual Framework

For this study, Andersen’s Behavioral Model of Health Services Use [17] will be used to guide the analysis of the literature review. This model identifies predisposing, enabling, and need factors as the key determinants of health service use [17]. This model is the best fit for this scoping review as it was conceptualized to explore factors that influence health service utilization, making it fit for studying access to contraceptive services in the population. Furthermore, the model also looked at disparities in access to health care, making it suitable for studying vulnerable populations such as low-income, underinsured, or uninsured women. Moreover, the model directly ties into access to healthcare and will be applied to structure and categorize the interventions based on how various interventions address these key determinants of contraceptive use among uninsured and underinsured women.

### Stage 1: Identifying the research question

This scoping review will aim to scope literature regarding the intervention strategies focused on improving access to contraceptive services for uninsured and underinsured women in the United States. “What intervention strategies have been implemented to improve access to contraceptive services for uninsured and underinsured women in the United States?”

Secondary objectives are to describe (i) intervention components, (ii) implementation contexts, and (iii) reported outcome metrics.

The Population, Concept and Context (PCC) framework for determining the eligibility of research questions has been applied to the study question, as shown in Table 1 below.

**Table 1.** PCC Application.

|  |  |
| --- | --- |
| Title: Bridging the Access Gaps in Contraceptive Services: A Scoping Review of Intervention Strategies for Uninsured and Underinsured Women in the USA |  |
| Primary Question | What intervention strategies have been implemented to improve access to contraceptive services for uninsured and underinsured women in the United States? |
| Population | Uninsured and underinsured women in the U.S. |
| Concept | Intervention strategies aimed at improving access to contraceptive services. |
| Context | United States healthcare, health departments and community settings, including policy, programs and service delivery environments. |

### Stage 2: Identifying relevant studies

Relevant studies will be identified by conducting a comprehensive search on PubMed, ScienceDirect, and Web of Science for peer-reviewed articles. Additional databases such as EMBASE, CINAHL, Scopus, and PsycINFO will be searched to improve coverage of public health and social science literature. Grey literature, such as project reports and conference abstracts on contraceptive programs/interventions/policy, will be retrieved from the websites of public health organizations, conferences, governmental agencies (e.g., CDC, Guttmacher Institute), and non-governmental organizations that focus on reproductive health. ProQuest Dissertations & Theses Global” and ClinicalTrials.gov will be searched for unpublished intervention evaluations. The reference lists of studies selected from the database search will also be screened for relevant articles to ensure they are included.

The search strategy will include relevant search and medical subject headings (MeSH) terms related to the population (e.g., “uninsured women,” “underinsured women,” “low-income women”), concept (e.g., “contraceptive access,” “reproductive health services,” “family planning”), and context (e.g., “United States,” “policy” “interventions,” “family planning programs,” “healthcare programs”). The expertise of an experienced subject librarian will be consulted to refine key terms and finalize the search strategy. Boolean operators, such as “AND” and “OR”, will be used to combine the search terms as shown in this preliminary Pubmed/MEDLINE Preliminary Search Strategy “(contraception OR “birth control” OR “family planning” OR “Intrauterine Device” OR condoms OR “barrier method” OR “tubal ligation”) AND (underinsured OR under-insured OR uninsured OR “public insurance” OR “no insurance” OR “insurance status”)”. The search strategy will be refined for each database as necessary.

The literature review will include only studies published in the English language due to resource constraints for translation and will consider studies published between January 2005 and June 2025. The period 2005-2025 was chosen because it captures the Affordable Care Act implementation and subsequent state-level policy changes influencing contraceptive access.

### Stage 3: Study selection

The identified studies will be scrutinized for duplicates, and duplicates will be removed. The study selection will be done in two steps. First, Title-and-abstract screening will be conducted independently by two reviewers using Rayyan; conflicts will be resolved through discussion or a third reviewer. Secondly, the full text of studies that pass the first step will be reviewed for eligibility by two authors separately, and discrepancies between the two reviewers will be discussed by a third reviewer. The eligibility criteria were designed to focus only on the articles that address issues described in the research question: “What intervention strategies have been implemented to improve access to contraceptive services for uninsured and underinsured women in the United States?” The inclusion criteria are;

1. Studies that present intervention strategies and were published in the English language between the years 2005 and 2025,
2. Studies that present intervention strategies that were published in the USA,
3. Studies that present intervention strategies on women aged 15–49 years,
4. Studies that present intervention strategies on outcomes related to contraceptive access,
5. Studies that present strategies for uninsured, underinsured, or low-income women,
6. Studies and grey literature that present intervention strategies aimed at improving access to contraceptive services for uninsured or underinsured women.

The exclusion criteria are;

1. Studies with no intervention strategies on access to contraceptives,
2. Studies exclusively involving fully insured populations will be excluded; mixed-population studies will be retained if data for uninsured/under-insured women are extractable,
3. Studies focused on interventions outside the U.S.

The preferred Reporting Items for Systematic Reviews and Meta-analyses extension for scoping review (PRISMA-ScR): Checklist and Explanation [18] will be employed to present the literature search and screening process result as shown in Figure 1 below;

**Figure 1.**
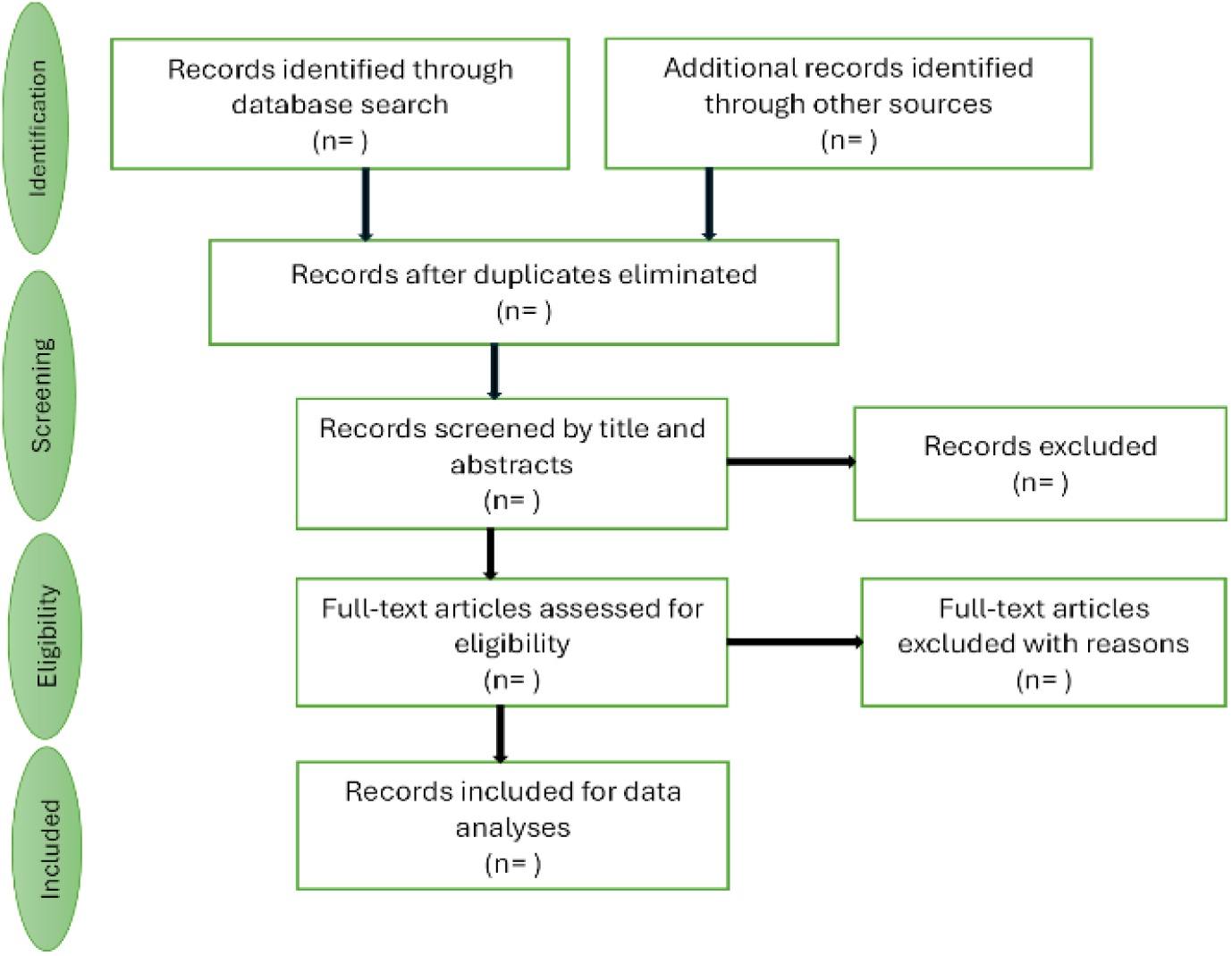
PRISMA flow chart demonstrating literature search and selection of studies.

### Stage 4: Charting the data

Data will be extracted and charted by one reviewer and checked for accuracy by a different reviewer using a piloted form in Excel; discrepancies will be reconciled by consensus. The Template for Intervention Description and Replication (TIDieR) checklist [19] for reporting interventions will be used to extract data from the selected intervention studies/programs. This will ensure that important components and standardized information are extracted from each selected study. The TIDieR checklist will be modified to include reference information (author and year of publication), methodology (i.e., study design), for whom (i.e., target population) and the model determinants, as shown in Table 2 below,

**Table 2.** Data charting domains and explanations adapted from the Template for Intervention Description and Replication (TIDieR) [19].

| Data Charting Domain | Explanation |
| --- | --- |
| Reference information | Name of author and year of publication |
| Brief name | Name or a phrase that describes the intervention |
| Intervention type | Description of the intervention type (e.g., policy change, community health program) |
| Why | Describe any rationale, theory, or goal of the elements essential to the intervention. |
| What | **Study design:** for example, quantitative study, qualitative study<br>**Materials:** describe any physical or informational materials used in the intervention, including those provided to participants or used in intervention |
|  | delivery or in the training of intervention providers. Provide information on where the materials can be accessed (such as online appendix, URL)<br>**Procedures:** describe each of the procedures, activities, and/or processes used in the intervention, including any enabling or support activities |
| Who provided | For each category of intervention provider (such as psychologist and nursing assistant), describe their expertise, background, and any specific training given. |
| For whom | Describe the target population/audience for which the intervention was designed. |
| How | Describe the modes of delivery (such as face-to-face or by some other mechanism, such as Internet or telephone) of the intervention and whether it was provided individually or in a group. |
| Where | Describe the type(s) of location(s) where the intervention occurred, including any necessary infrastructure or relevant features. |
| When and how much | Describe the number of times the intervention was delivered and over what period of time, including the number of sessions, their schedule, and their duration, intensity, or dose. |
| Tailoring | Tailoring If the intervention was planned to be personalized, titrated, or adapted, then describe what, why, when, and how |
| Modifications | If the intervention was modified during the course of the study, describe the changes (what, why, when, and how) |
| How well | **Planned:** If intervention adherence or fidelity was assessed, describe how and by whom, and if any strategies were used to maintain or improve fidelity, describe them.<br>**Actual:** If intervention adherence or fidelity was assessed, describe the extent to which the intervention was delivered as planned. |
| Model determinant addressed | If and which of the factors of Andersen's Behavioral Model of Health Services Use was addressed (e.g. predisposing/enabling/need). |

### Stage 5: Collating, summarizing, and reporting the results

The data extracted from each study will be collated, summarized, and reported descriptively and thematically using narrative synthesis. The descriptive summary will provide a numerical summary of the types of studies included, the geographic distribution of studies, and the type of interventions implemented. We will present a visual evidence map aligning intervention types with PCC elements and a frequency table of outcomes measured.

Thematic analysis will be conducted to identify common themes across the studies. This will include analyzing how different types of interventions address the predisposing, enabling, and need factors identified in Andersen’s Behavioral Model. Furthermore, narrative synthesis will be used to synthesize, summarize and explain the overall findings of the studies included.

## Discussion, ethics and dissemination

This paper presents the protocol for a scoping review of intervention strategies aimed at improving access to contraceptive services for uninsured and underinsured women in the United States. We expect to identify strategies, programs, and policy-related interventions, as well as gaps in current knowledge, among this population. Based on the findings from this review, recommendations will be made for future research, program development, and policy. Furthermore, a potential limitation is that the scoping review will not assess the methodological quality of included studies, as such findings will map the breadth of existing evidence but will not draw conclusions about the comparative effectiveness of interventions. In addition, the review will be limited to studies published in English and those accessible through selected databases and grey literature sources, which may result in the exclusion of potentially relevant data from non-English contexts or unindexed sources.

Moreover, as the review uses exclusively publicly available data, it is exempt from Institutional Review Board oversight. The results of the review will be disseminated via a peer-reviewed journal and presented at national conferences. Finally, any amendments to the protocol, including changes to eligibility criteria, search strategy, or data extraction methods, will be transparently documented and updated in the registered protocol on the Open Science Framework. If unforeseen challenges arise that significantly affect the feasibility or relevance of the review, the research team will reassess the scope or consider protocol termination, with justifications and updates recorded publicly to ensure transparency and accountability.

## Data Availability

No datasets were generated or analysed during the current study. All relevant data from this study will be made available upon study completion.

## Supporting information

S1 File. PRISMA 2020 checklist.

(DOCX)

